# An Unsupervised Approach to Identify Patient-Specific EMG Detector to Trigger Robot-Assisted Therapy

**DOI:** 10.1101/2024.12.06.24318597

**Authors:** Monisha Yuvaraj, A.T. Prabakar, Varadhan S.K.M, Etienne Burdet, Ander Ramos-Murgialday, Sivakumar Balasubramanian

## Abstract

In severely impaired stroke patients, implementing EMG-driven robot-assisted therapy requires the presence of sufficient residual EMG and a patient-specific detector for accurate and low-latency EMG detection. However, identifying such a detector is challenging, especially when the level of residual EMG in a given patient is unknown . This paper proposes an unsupervised approach to distinguish between EMG data when the patient is relaxed versus attempting a movement – the *maximally separating detector*. We investigated six different detector types and separation measures using EMG data from a previous randomized controlled trial. The results indicate that the approximate generalized likelihood ratio detector, along with the modified Hodges and modified Lidierth detectors, achieved the best separation. Using a subset of clinician annotated data to evaluate the detection performance, the modified Hodges detector employing the probability difference-sum ratio measure had the best detection performance in terms of detection accuracy and latency. Using the data from 30 participants, we propose a probability difference-sum ratio threshold of 0.7 for the modified Hodges detector to identify patients with sufficient residual EMG to trigger robotic assistance. From the results, we propose the use of modified Hodges detector along with a probability difference-sum ratio measure to learn the maximally separating detector for a given patient, which will screen the patient for sufficient residual EMG and provide a detector to trigger robotic assistance if sufficient EMG is present. The validation of this approach using a large dataset and investigating the quality of the human-machine interaction implemented with such a detector is warranted.

## 1 Introduction

Stroke affects about 12.2 million people around the globe every year, and hand impairments are commonly seen after a stroke (1). Robot assisted therapy facilitates high-intensity movement training, which enhances sensory-motor recovery after a stroke (2). A robot typically provides active-assistance by offering the appropriate assistance to maximize the patient’s active participation in completing movements. Active participation is essential for recovery as no improvement in clinical scales were observed with passive therapy that does not involve voluntary participation from the patient (3). To ensure patient’s maximum active participation, the active-assisted training mode uses patient’s residual movements to identify their movement intention to titrate robotic assistance. Too much assistance can lead to slacking (4), too little can affect motivation (5), while poorly timed assistance can affect the sense of agency (5).

About 30% of stroke patients are severely impaired (6), with no residual movements; detecting movement intention in this patient population is a challenge . Electroencephalogram (EEG)-based brain computer interface (BCI) is commonly employed to detect movement intent in this population for assisted therapy (7). However, our recent work has shown that electromyography (EMG) from target muscles is a viable alternative for EEG-BCI, as EMG has better signal to noise ratio (SNR), is task specific and easy to set-up for routine use (8). In this context ensuring a naturalistic human-machine interaction to detect the intention to move is vital for maximizing the patient’s active participation, while maintaining their motivation and the sense of agency. For EMG-based robot assisted therapy this means that, the robot should provide assistance only when there is movement intention (i.e., minimal false positives and false negatives), and quickly with minimal detection latency. This requires an accurate and low-latency EMG detector for the closed-loop control of robotic assistance. Several EMG onset time detectors have been proposed (9; 10; 11; 12; 13), with no standardized method or a systematic way to determine the best detector for EMG-triggered robot assisted therapy. In our previous study (14), we systematically investigated the performance of various EMG detectors in the literature using simulated low SNR EMG signals. We identified the simple threshold-based modified Hodges detector, the statistical decision-based approximate generalized likelihood ratio (AGLR) detector, and the fuzzy entropy detector as the best performing detectors in terms of detection accuracy and latency. This identification was possible because the ground truth about the presence/absence of the simulated EMG data was fully known. Although this “supervised” approach provided an understanding of the performance of existing detectors on low SNR EMG signals, it is unsuitable for identifying patient-specific optimal EMG detectors using real signal recordings from target muscles of severely impaired patients. As, with real muscle recordings, there is no ground truth about the presence/absence of EMG activity, there is a need for an unsupervised approach to identify the optimal detector when working with real data from severely affected stroke patients who exhibit no visible movement.

Therefore this paper presents an unsupervised technique to (a) screen severely impaired patients for the presence of sufficient residual EMG that can be used for EMG-triggered robot assisted therapy, and (b) identify the best detector parameters for accurate and fast pick-up of movement intention from this residual EMG signal of patients that have sufficient residual EMG. The presented method is tested with the EMG data collected from a previously published randomized controlled trial evaluating the efficacy of robot-assisted therapy contingent on movement intention detected through event-related desynchronization of EEG (7).

## 2 Methods

The EMG data were collected from thirty-two chronic stroke participants with a cFMA (combined arm and hand modified FMA) score of 12.15 ± 8.8, who could not actively extend their wrist and fingers. Two patients were excluded from the original study due to equipment malfunction during training (7). Surface EMG was recorded during all robot-assisted upper limb therapy sessions from four upper-limb muscles: *extensor carpi ulnaris, extensor digitorum, biceps*, and *triceps*. Details about patient demographics and the intervention can be found in (7). EMG data from the *extensor carpi ulnaris* and *extensor digitorum* muscles of the paretic side were used as individual trials for the purpose of this study.

During the intervention, in each trial, the rest phase was 3 seconds long, where the subjects were given the instruction about the movement to be made, followed by a “READY” and a “GO” cue, separated by 2 seconds and 1 second, respectively. Following the “GO” cue, the subjects were asked to perform the desired movement for 5 or 6 seconds, resulting in a total duration of 11-12 seconds for each trial. In the current analysis, the rest period (RP) for a trial is defined as the first 4 seconds of a trial, and the move period (MP) is defined as the remaining 7-8 seconds of the trial Fig.2.1 (a)).

The EMG signals were sampled at 500 Hz. Let *x*_*t*_ [*n*] represent the recorded muscle activity for a given trial *t*, where *n* is the sampling instant. The total number of samples in each trial *x*_*t*_[*n*] is *N*_*t*_ ∈ {5500, 6000}, where the rest phase is defined between the interval 0 ≤ *n < N*_*r*_, with *N*_*r*_ = 2000. Fig.2.1 (a) explains the structure of each trial of EMG data.

### 2.1 How to identify the best EMG detector without a ground truth signal?

An EMG detector maps the raw EMG data *x*_*t*_ [*n*] from the trial *t* to a binary output *y*_*t*_ [*n*], which is 1 when muscle activity is present and 0 otherwise.

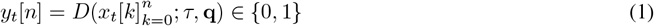

where, *D* (·) is the detector function, *τ* ∈ ℝ is the threshold parameter to convert the detector’s test function into a binary output (Fig. 2.1), and 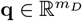 is the set of *m*_*D*_ hyper-parameters of the detector. The parameter *q* depends on the nature of the detector and changes from one detector type to other. For instance, **q** = {*α, f*_*c*_ }for modified Hodges detector where *α* is the weight for the threshold and *f*_*c*_ is the cut-off frequency of the low pass filter. For the AGLR detector **q** = {*α, W*}; where *α* is the weight for the threshold, and *W* is the window size in ms for computing the test function.

In the current analysis, the threshold *τ* is estimated for a trial using the data from the rest period ignoring the first 1 second of the data 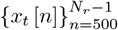, as follows:

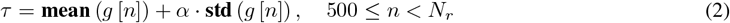

where, *g*[*n*] is the test function computed by the detector using the input signal *x*[*n*]. This threshold *τ* is then used to detect the presence of EMG in the data from the move period of the trial 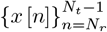 . The detector output *y*_*t*_ [*n*] for trial *t* is given by the following:

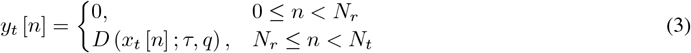

Six different detector types were investigated in the current study, *D*_1_ to *D*_6_, listed in Table 1 along with their test functions and parameters. The six detectors include, the Fuzzy Entropy (*D*_1_), Root mean square (RMS) detector (*D*_2_), Modified Lidierth (*D*_3_), Modified Hodges (*D*_4_), AGLR-G (*D*_5_) and AGLR-L (*D*_6_).

**Table 1:** Description of the EMG detectors.

| Labels | Detectors | Test functions | Parameters |
| --- | --- | --- | --- |
| D1 | Fuzzy Entropy | Chebashyv distance<br>Fuzzy function | Window size (W)<br>Weight ($\alpha$) |
| D2 | RMS | Root Mean Squared | Window size (W)<br>Weight ($\alpha$)<br>Window shift (p)<br>Temporal threshold (m) |
| D3 | Modified Lidierth | Rectified<br>Low pass filtered | Cut-off frequency ($f_c$)<br>Weight($\alpha$)<br>Double threshold params |
| D4 | Modified Hodges | Rectified<br>Low pass filtered | Cut-off frequency ($f_c$)<br>Weight($\alpha$) |
| D5 | AGLR-G | Likelihood ratio test<br>(Gaussian Dist) | Window size (W)<br>Weight($\alpha$) |
| D6 | AGLR-L | Likelihood ratio test<br>(Laplacian Dist) | Window size (W)<br>Weight($\alpha$) |

The performance of the given detector with hyper-parameters *q* can be evaluated if we had access to the ground truth about the presence/absence of EMG in the move period of the given trial. This can be done by computing the detection accuracy and latency, as was done in our previous work (14). How to evaluate the detector’s performance when the ground truth is unavailable? One approach is to compare the output of the detector on data from two conditions: (a) when the patient is relaxed, where we can safely assume the data to be devoid of any EMG activity; we refer to this as the *null hypothesis* ℋ_0_, and (b) when the patient attempts to move, where there might be residual EMG activity if the patient has spared cortical connections to the target muscle; we refer to this as the *alternative hypothesis* ℋ_1_. We use the proportion of 1s in the detector’s output as a measure of its ability to detect EMG or the probability of detecting EMG in a given data. Then, we would expect the following from a good detector:

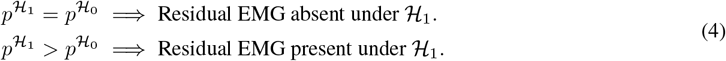

where, 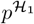and 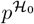 are the detection probabilities under ℋ_1_ and ℋ_0_; note that the equality and inequality signs used here are in the statistical sense. If a given participant does not have residual EMG in his/her target muscles, then 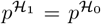 will be true for any detector. On the other hand, if residual EMG is present, then 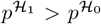 for appropriately designed detectors. The best detector would be the one providing the maximal separation between data under ℋ_1_ and ℋ_0_ for some appropriate measure of separation. We define such a detector as the *maximally separating detector* for given separation measure.

### 2.2 Data preparation under ℋ_0_ and ℋ_1_

We require data generated independently under the ℋ_0_ and alternate ℋ_1_ to employ the method described above. Note that the rest period data cannot be used because it is already used by the detector to estimate the threshold *τ* for the test function for each trial. To address this issue, we generated artificial data 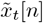 for each trial such that: (a) in the rest period 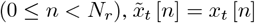 and (b) in the move period 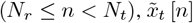 has the same first (mean) and second order (autocovariance) moments as that of the rest period data. The procedure for generating 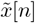 is explained in Algorithm 1; where, 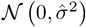 represents a normal distribution with zero mean and variance 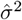.

#### Algorithm 1

To generate 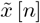.

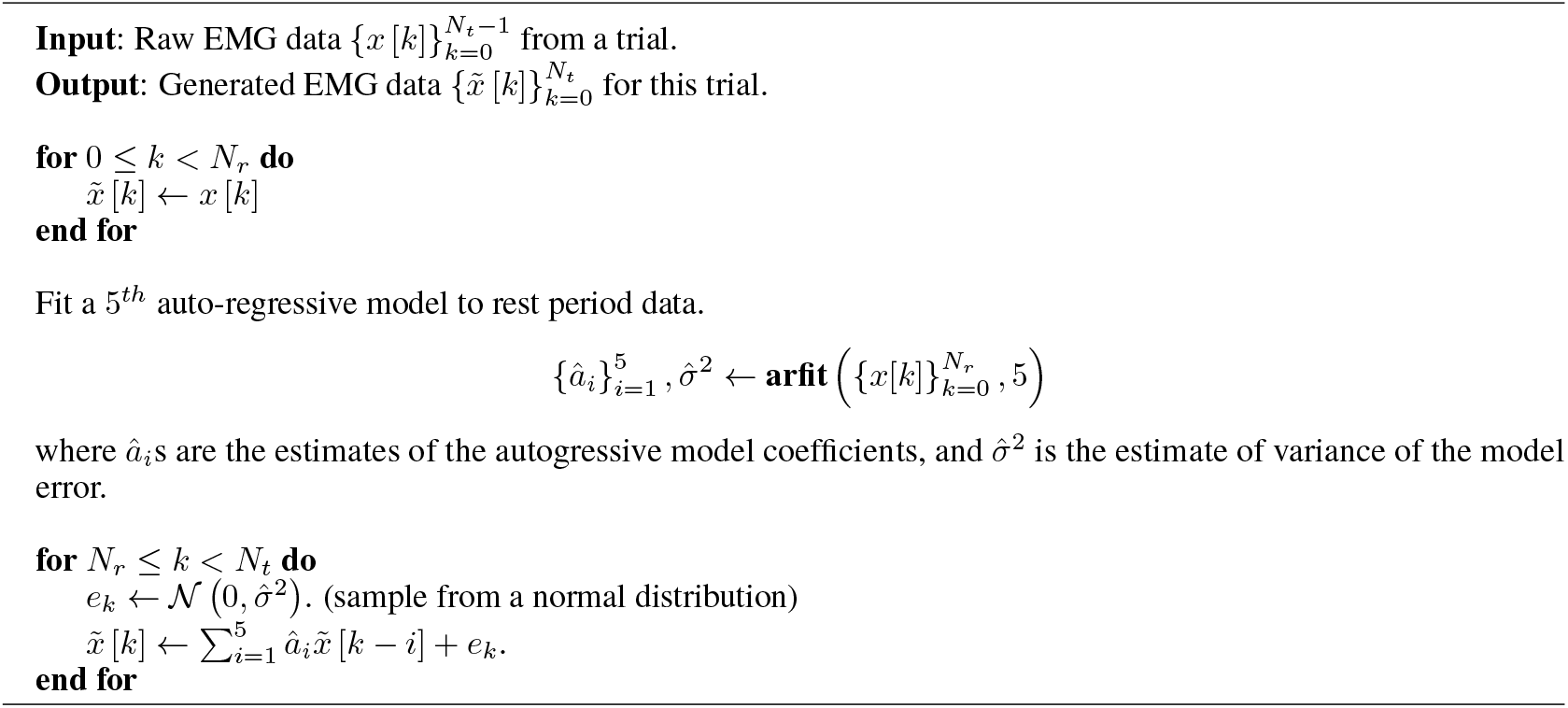

In the EMG dataset used in the current study, some trials with large repeated spikes were identified. Due to the presence of these spikes the AR model overestimates the variance of the error while generating the move period data of the trials under ℋ_0_. This would lead to a case where 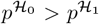, causing incorrect parameter optimization. Therefore these trials were removed by computing the total variation distance (TVD) between the rest period and move period of the trials under ℋ_0_. The trials with a very high TVD values, which indicates dissimilarity between the two periods were removed along with the corresponding trial under ℋ_1_.

Following the removal of these trials for all 30 subjects, the dataset had for each trial *t* of a given subject, (a) the actual recorded signals *x*_*s*,*t*_ [*n*] when the patient attempted movements, i.e., data generated under ℋ_1_, and (b) the artificial data 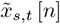 generated using the autoregressive model, which is data from ℋ_0_. Both data are passed through a given detector *D* to obtain its output 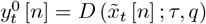 and 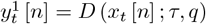, which are then used to compute the detection probability under ℋ_0_ and ℋ_1_ for each movement trial.

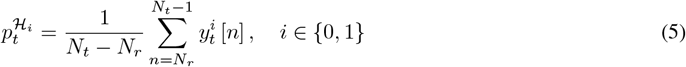

where, 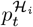 is the EMG detection probability for the trial *t* under ℋ_*i*_.

Let *T*_*s*_ be the number of movement trials performed by a given subject *s*, with 1 ≤ *s* ≤ 30. Thus, for subject *s*, we have a set of *T*_*s*_ estimates of 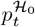 and 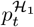 with 1 ≤ *t* ≤ *T*_*s*_,

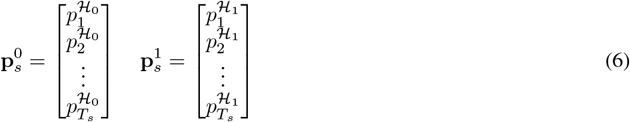

The detection probabilities from across all trials, 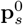 and 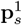, can be used to determine: (a) if the subject *s* has residual EMG, and (b) if there is residual EMG, what is the best detector to separate the recorded data under the rest and move periods. This requires the choice of a separation measure to compare the probabilities of detection under ℋ_0_ and ℋ_1_.

### 2.3. Separation measures to compare ℋ_0_ and ℋ_1_

There are multiple ways to define a measure of separation between the given estimates of detection probability under ℋ_0_ and ℋ_1_. Since, the choice of the separation measure can impact the outcomes, we explored six different separation measures: three variations of the total variation distance (TVD), difference in probabilities (DP), likelihood ratio (LR), and the probability difference-sum ratio (PDSR).

**The Total Variation Distance (TVD)** is a measure of the difference between two probability density function *f*_1_ (*x*) and *f*_2_ (*x*) defined as

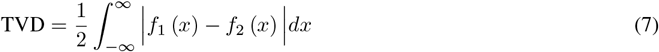

We can compute the TVD from 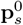 and 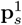 by first computing their corresponding histograms, from which we can compute the TVD using Eq. 7. Since the histogram estimates and the TVD were sensitive to the number of histogram bins, we computed the TVD for three different number of bins - 10, 20, and 100, which are referred to as *S*_1_, *S*_2_ and *S*_3_ measures, respectively.

While the TVD provides a single number to quantify the separation between the detection probabilities under ℋ_0_ and ℋ_1_, the following three measure work with the detection probabilities from individual trials to quantify separation. The overall separation for each of the following measures are computed as the median of the separation measures across the different trials.

**Difference in probabilities (DP)** measures the amount by which the detection probability under ℋ_1_ is higher than that of ℋ_0_. It is defined as

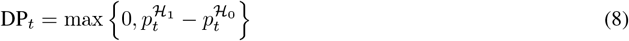

where, DP_*t*_ is separation for the trial *t*. The overall separation measure for DP is defined as 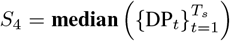 .

**Likelihood ratio (LR)** is the ratio of the probabilities under the ℋ_0_ and ℋ_1_, defined as

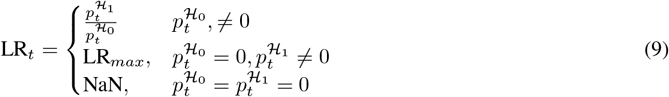

where LR_*t*_ is the separation measure for trial *t* and all trials with LR_*t*_ = inf was replaced with LR_*max*_ which is defined as 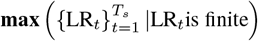, computed after obtaining LR_*t*_ for all trials. The overall separation measure for LR is defined as 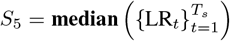.

**Probability Difference-Sum Ratio (PDSR)** is the combination of DP and LR, defined as

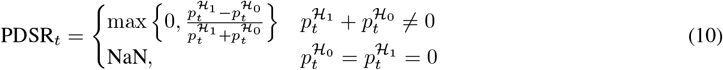

where PDSR_*t*_ is the separation measure for trial t. The overall separation measure for PDSR is defined as 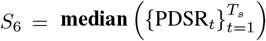. In this paper, we used the terms DP, LR, and PDSR synonymously with the overall separation measure *S*_4_, *S*_5_ and *S*_6_, respectively.

### 2.4 Maximally Separating Detectors: Patient specific optimisation of detectors

Given the uncertainty in the presence of residual EMG and its SNR in severely impaired stroke subjects, patient-specific detectors are warranted. To this end, we investigated six different detector types (*D*_1_ to *D*_6_) for each subject. The optimal parameters for each of these six detector types were identified as the ones that maximize the separation between the data under ℋ_0_ and ℋ_1_; the six different separation measures (*S*_1_ to *S*_6_) were employed for this purpose. For a given subject *s*, detector type *D*_*i*_, and separation measure *S*_*j*_, we determine the best parameter 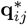 as the one that maximizes the value of the separation measure *S*_*j*_,

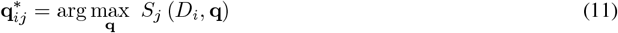

where, the value of *S*_*j*_ (•) depends on the detector type *D*_*i*_ and its parameters **q**. The detector with the optimal parameters 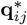 is the *maximally separating detector* of detector type *D*_*i*_ using the separation measure *S*_*j*_.

A brute-force grid search algorithm was employed to identify the different maximally separating detectors for the six detector types and six separation measures. For a given subject *s*, detector *D*_*i*_, and separation measure *S*_*j*_, the detector parameter values **q** were fixed and the detector’s output was computed. This was followed by the computation of the separation value *S*_*j*_ (*D*_*i*_, **q**). This procedure was repeated for all possible values of the detector parameters, and the best parameter combination was chosen as 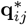. Repeating this procedure across the different separation measure and detectors resulted in a 6 *×* 6 grid of optimal detector parameters (or maximally separating detectors) for each subject.

### 2.5 How good is a maximally separating detector?

A maximally separating detector is only as good as its ability to accurately and quickly pick-up true muscle activity. It is *a priori* unclear which of the 6 × 6 maximally separating detector for a given subject will have maximum detection accuracy and minimum latency. It might not be possible to theoretically compute the detection accuracy and latency for a given detector type and a separation measure. Therefore, this needs to be determined empirically using real data. To this end, we computed the detection accuracy and latency using the data from the individual patients. For this purpose, we randomly selected five trials per session for each patient with a maximum of 50 trials per patient (the number of sessions per patient varying between 10 and 15). This exclusive set of 1500 trials – referred to as the *ground truth dataset* (GTD) – was not part of the unsupervised learning dataset. An expert neurologist (ATP with 11 years of experience with clinical EMG) marked the onset and offset of EMG in each trial of the GTD through visual inspection using a custom-built Matlab-based graphical user interface.

The ground truth labelled by the clinician was used for computing the “cost” of detection

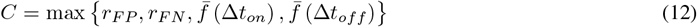

The *false positive* (*r*_*F P*_) is defined as the number of ‘1’s in the move period where there is no muscle activity which is computed as

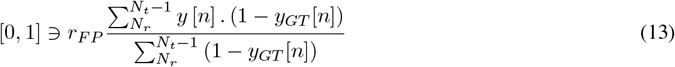

where *y*[*n*] is the binary output of the detector for a trial in GTD and *y*_*GT*_ [*n*] the ground truth of the same trial labelled by the clinician. The *false negative* (*r*_*F N*_) is defined as the number of ’0’s when the muscle is active, computed as the following.

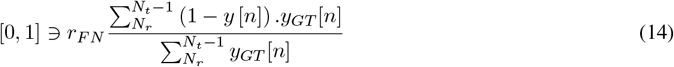

**Figure 2.1:**
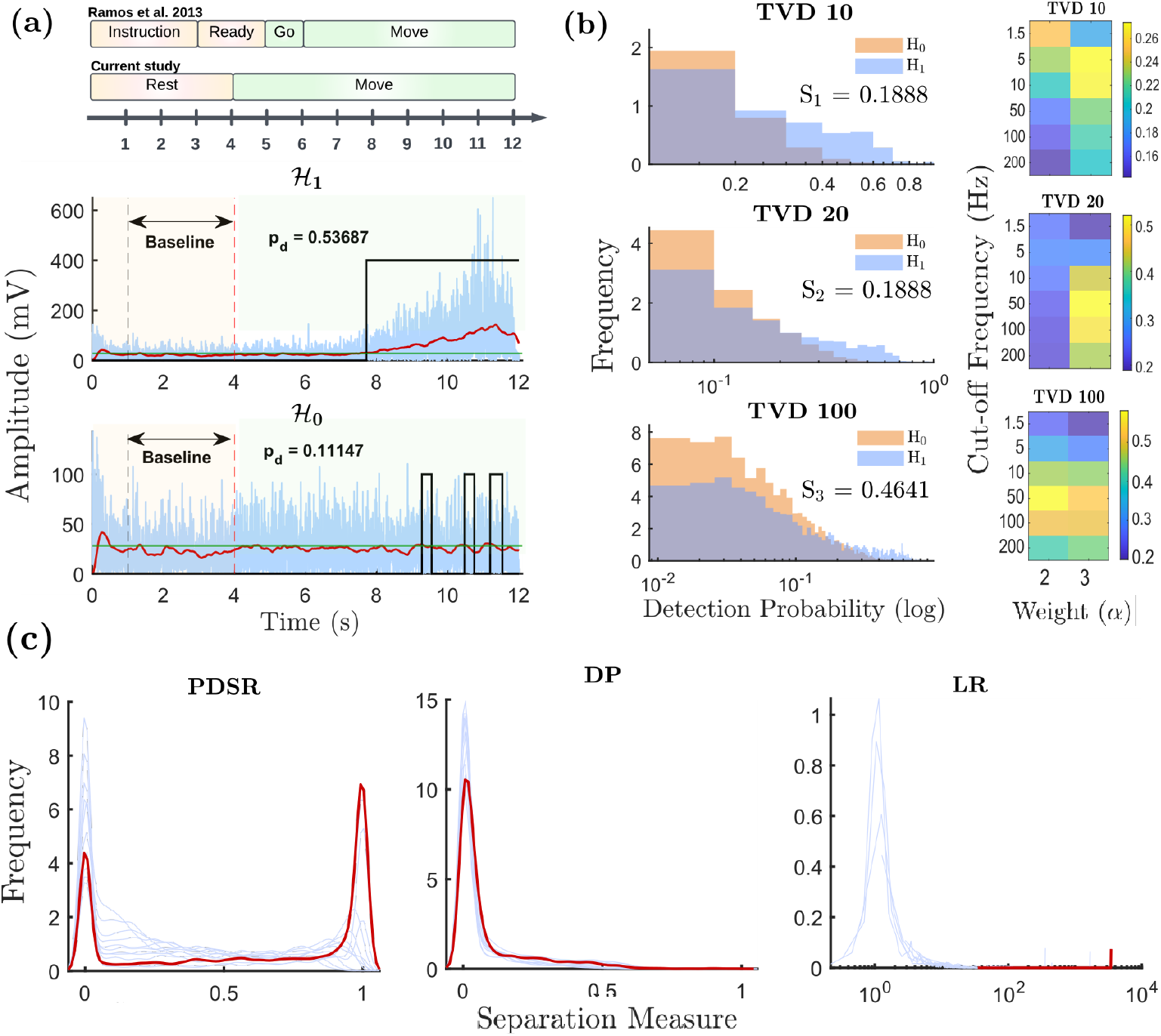
Optimizing the parameters of the detector using the unsupervised approaches. The figure presents the output of the modified Hodges detector of trial from one patient. (a) Trial structure of EMG as described in the previous and the current study is presented (above) ; Output of the modified Hodges detector (rectified EMG (blue), Binary output (black), low pass filtered signal (red), Threshold (green)) along with detection probability computed for both ℋ_0_ and ℋ_1_ trials (below) . (b) Left : Histogram of detection probability under ℋ_0_ and ℋ_1_ (TVD 10 & TVD 20 : Weight = 2, f _*c*_ = 1.5 Hz; TVD 100 : Weight = 3 and f_*c*_ = 200 Hz); Right : The heatmap of the TVD for the different bin numbers is presented, yellow box corresponds to the parameter with maximum TVD value for the modified Hodges detector. (c) The distribution of PDSR separation measure computed over different parameter combination (left). The distribution of DP separation measure computed over different parameter combination (center). The distribution of LR separation measure computed over different parameter combination (right; x axis in log scale for better visualization). Red curve corresponds to the distribution with the maximum median chosen as the *maximally separating detector*.

The move period of any trial will consist of time segments where there are and there aren’t muscle activity. The segments with muscle activity will be referred to as *active segments* (where *y*_*GT*_ [*n*] = 1), while the others will be referred to as *inactive segments* (where *y*_*GT*_ [*n*] = 0). Let there be *K* active segments in trial and there will be *K* or *K* − 1 inactive segments, depending on whether or not the given trial ends in an inactive segment or not, respectively. Note that here, we ignore the data before the first active segment in the move period.

Let 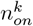 and 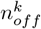 be start and stop times, respectively, of the *k*^*th*^ active segment, such that the following ordering is satisfied for all 0 ≤ *k < K*,

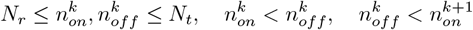

If the EMG detector’s output is 1 at least once during the *k*^*th*^ active segment, i.e., 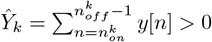 then onset detection latency is defined as

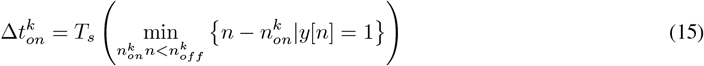

If *Ŷ*_*k*_ = 0, then 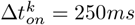. Similarly the offset latency of the *k*^*th*^ active segment is computed as follows only if the EMG detector’s output is 0 at least once during the inactive segment following the *k*^*th*^ active segment,

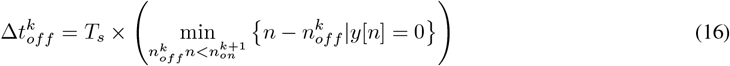

If the detector’s output never becomes 0 during the inactive segment, then 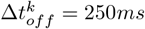.

The cost due to individual offset and onset latencies is quantified by separate functions 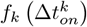 and 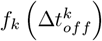 that maps all latencies above 250ms to 1. Further the average onset and offset latency for each trial is computed as follows:

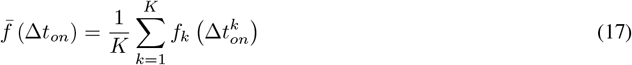

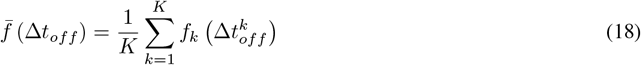

Note that for simplicity we have assumed the number of inactive segments to be *K* in Eq. 18. The detection cost was computed for all maximally separating detectors for all subjects, to identify the best detector type and separation measure that results in the maximally separating detector with the least detection cost.

### 2.6 Statistics Analysis

The performance of the detector types (Fig.3.1) was compared through individual linear mixed effect models for each separation measure to identify the detector with the maximum separation. Post-hoc Tukey’s Honest Significant Difference (HSD) test was performed to evaluate the pairwise differences among the detector types for each separation measure; the statistical significance was set at p *<* 0.0033 (0.05*/*15).

The effect of the detector type and separation measure on the detection cost was investigated using another linear mixed effects model with the two fixed effect (detector type and separation measure) and one random effect (subjects). Pairwise comparisons were carried out to determine the best detector type across separation measures, and the best separation measure across detector types. Post-hoc Tukey’s HSD pairwise comparison of the detection cost was performed individually for the separation measures and for the detector types where the statistical significance was set at *p <* 0.0033 for both the comparisons. Significance of random effect was determined by comparing the mixed-effects model with an ordinary least squares (OLS) model (without considering the random effects), using a Likelihood Ratio Test. Furthermore, another pairwise multiple comparison was performed to identify the best detector-separation measure pair that incurs the least cost (Fig.3.2); statistical significance was set at *p <* 0.000079(0.05*/*630).

All the analysis presented in this paper were carried out in MATLAB R2023, except for the statistical analysis. The linear mixed-effects modeling was performed using the ‘statsmodels’ package in python [RRID:SCR008394].

**Figure 3.1:**
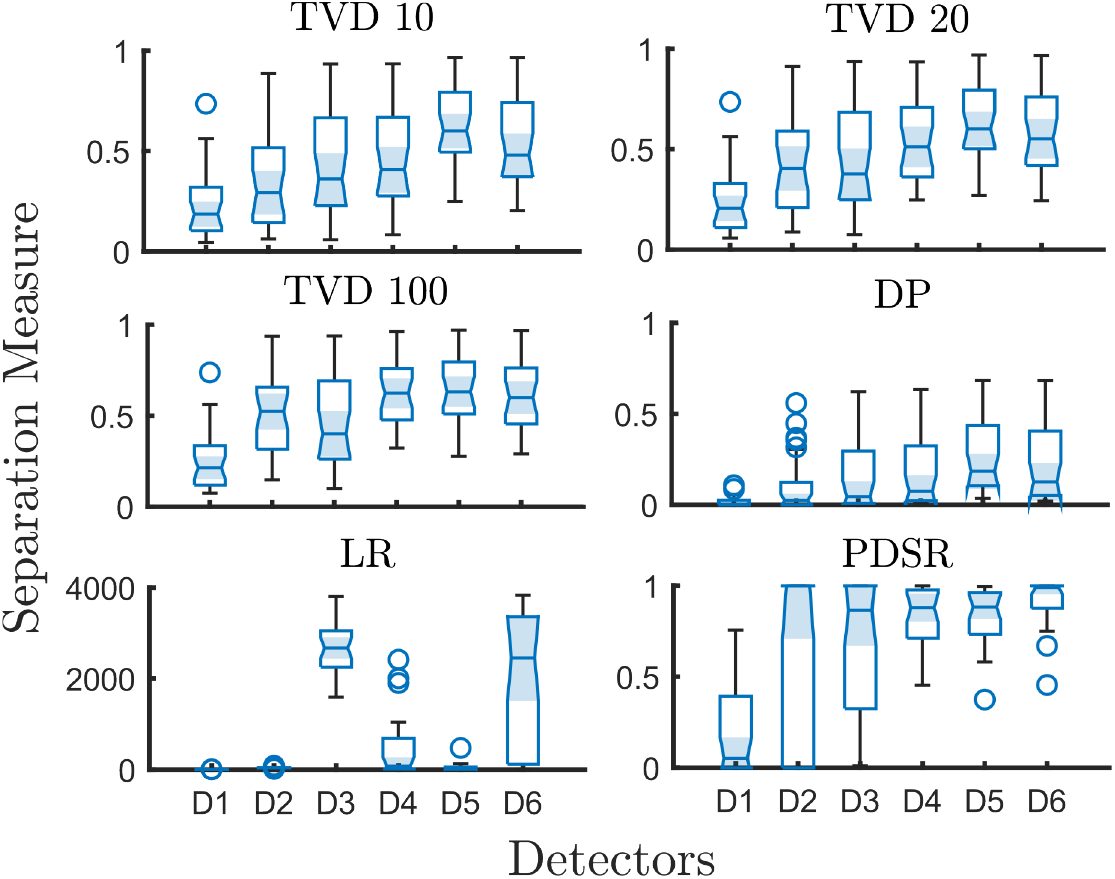
Box plots comparing the maximally separating detectors optimized using the six separation measures is presented. Each box represents data from all the patients. **D1**:Fuzzy Entropy; **D2**:RMS; **D3**:Modified Lidierth; **D4**: Modified Hodges; **D5**: AGLR-G; **D6**: AGLR-L

## 3 Results

A representative plot of the EMG data from a trial of a patient analysed using the modified Hodges detector is shown in Fig. 2.1 (a). The plot of the rectified EMG data (blue trace) and the lowpass filtered version (red trace) of this signal under ℋ_0_ and ℋ_1_ are depicted in the bottom plot of Fig. 2.1 (a). The green trace is the threshold *τ* estimated for the trial using the baseline data. The black trace is the output of the modified Hodges detector for the data. The plot of the histograms of the probability of detection 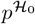 and 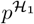 for this subject for the modified Hodges detector across multiple trials are shown in Fig. 2.1 (b) for the three different bin numbers (10, 20 and 100). The heatmap of the TVD computed for the different bin numbers using the histograms are shown next to these histograms as a function of the two parameters of the modified Hodges detector – cut off frequency and weight. The kernel density estimates of the other three separation measures, PDSR, DP, and LR, are shown in Fig.2.1 (c). The individual blue traces are the kernel density estimates across the different trials for a given detector parameter combination. The red trace is the one for the optimal detector parameters for the corresponding separation measure.

### 3.1 How well do the detectors distinguish between the rest and the move phase?

The patient-specific maximally separating detector was determined using Eq. 11 for the 6 separation measures and the 6 detector types; this results in a patient-specific 6 × 6 grid of maximally separating detectors. These detectors were compared to identify the one that gives the best separation between the rest and move phase data for the different separation measures. Fig.3.1 presents a box plot of the separation measure values for the 6 different maximally separating detectors for each separation measure. An individual linear mixed effects model for each separation measure revealed significant main effects of the detector type (*p <* 0.001). Pairwise comparison indicated that the statistical decision-based detectors (AGLR-G and AGLR-L), followed by the threshold-based detector (Modified Hodges and Modified Lidierth), achieved the highest separation across all the separation measures, except for the LR separation measure (Fig. 3.1); AGLR-L and the modified Lidierth detector have higher separation with LR separation measure.

### 3.2 What is the detection cost of the different maximally separating detectors?

The effect of the detector types and the separation measure on the detection cost of the different maximally separating detectors was evaluated using a linear mixed effects model. There was significant main fixed effect of the detector types (coeff = 0.037; *p <* 0.001) and the separation measure (coeff = 0.005; *p <* 0.01) on the detection cost. There was significant random effect (*p* = 0; *χ*^2^ = 1269.75). Post-hoc multiple comparison of the different separation measures reveal that the separation measure PDSR incurs the least cost across all detectors types, and the modified Hodges detector incurs the least cost across the different separation measure. Table 2 shows the mean difference in cost between the different pairs of separation measures; a negative mean differences implies that group2 has a lower detection cost than group1. In Table 2, PDSR has the most negative mean difference, that is statistically different from the other detectors, indicating that it incurs the least detection cost (highlighted bold text in Table 2). Similarly in Table 3, the modified Hodges detector incurs the least cost. Note that the detection cost of modified Lidierth is not statistically different from modified Hodges.

**Table 2:** Post-hoc analysis following a linear mixed effects model: Tuskey HD multiple comparison of the different separation measures. Mean difference = Group2 - Group1; therefore a negative mean difference implies the group2 has a mean lower than group1.^*\**^ indicates *p <* 0.05

| Group 1 | Group 2 | Mean difference |
| --- | --- | --- |
| TVD 10 | TVD 20 | -0.0155 |
| TVD 10 | TVD 100 | -0.0017 |
| TVD 10 | DP | 0.0197* |
| TVD 10 | LR | -0.0118 |
| TVD 10 | PDSR | **-0.0738\*** |
| TVD20 | TVD 100 | 0.0138 |
| TVD20 | DP | 0.0352* |
| TVD20 | LR | 0.0037 |
| TVD20 | PDSR | **-0.0583\*** |
| TVD100 | DP | 0.0213* |
| TVD100 | LR | -0.0101 |
| TVD100 | PDSR | **-0.0721\*** |
| DP | LR | -0.0315 |
| DP | PDSR | **-0.0935\*** |
| LR | PDSR | **-0.062\*** |

**Table 3:** Post-hoc analysis following a linear mixed effects model: Tuskey HD multiple comparison of the different detector types.^*\**^ indicates *p <* 0.05

| Group 1 | Group 2 | Mean difference |
| --- | --- | --- |
| AGLR-G | Fuzzy Entropy | 0.1374* |
| AGLR-G | RMS | 0.3003* |
| AGLR-G | AGLR-L | 0.3956* |
| AGLR-G | Modified Lidieth | -0.0949* |
| AGLR-G | Modified Hodges | **-0.0961\*** |
| Fuzzy Entropy | RMS | 0.163* |
| Fuzzy Entropy | AGLR-L | 0.2582* |
| Fuzzy Entropy | Modified Lidieth | -0.2323* |
| Fuzzy Entropy | Modified Hodges | **-0.2334\*** |
| RMS | AGLR-L | 0.0952* |
| RMS | Modified Lidieth | -0.3953* |
| RMS | Modified Hodges | **-0.3964\*** |
| AGLR-L | Modified Lidieth | -0.4905* |
| AGLR-L | Modified Hodges | **-0.4916\*** |
| Modified Lidieth | Modified Hodges | -0.0011 |

Fig.3.2 shows the detection cost of the 36 detector-separation measure pairs depicted in the decreasing order of their corresponding mean detection cost. In general, the modified Hodges detector has the least cost across the different separation measures, with the modified Hodges detection and PDSR separation measure pair (MH-PDSR) incurring the lowest mean detection cost. Cost of the modified Lidierth-PDSR pair is also statistically not different from the MH-PDSR pair. The modified Hodges detector perform consistently well in combination with all the separation measure except for the DP separation measure.

### 3.3 PDSR separation measure – a screening tool for the presence of residual EMG

The value of a separation measure computed on EMG data using an optimal detector is an indicator of the presence of residual EMG, which could be used to screen patients for the presence/absence of residual EMG for robot-assisted therapy. To this end, we depict the kernel density plot of the PDSR separation measure obtained using the *maximally separating detector* of the modified Hodges detector type for the 30 stroke participants in the current study (Fig.3.3 (b)). In Fig.3.3 (a) we present the scatter plot between the PDSR and the mean probability of detection under ℋ_1_; these two variables are positively correlated with the Spearman correlation coefficient of 0.721. Based on these two plots, we propose a threshold of 0.7 for the PDSR of a maximally separating modified Hodges detector for screening patients for the presence/absence of residual EMG. Below this threshold, the probability of detection under ℋ_1_ is close to 0, and it rises sharply beyond this threshold. This indicates that patients with PDSR above 0.7 are likely to consistently produce robot-assisted movements using their residual EMG.

**Figure 3.2:**
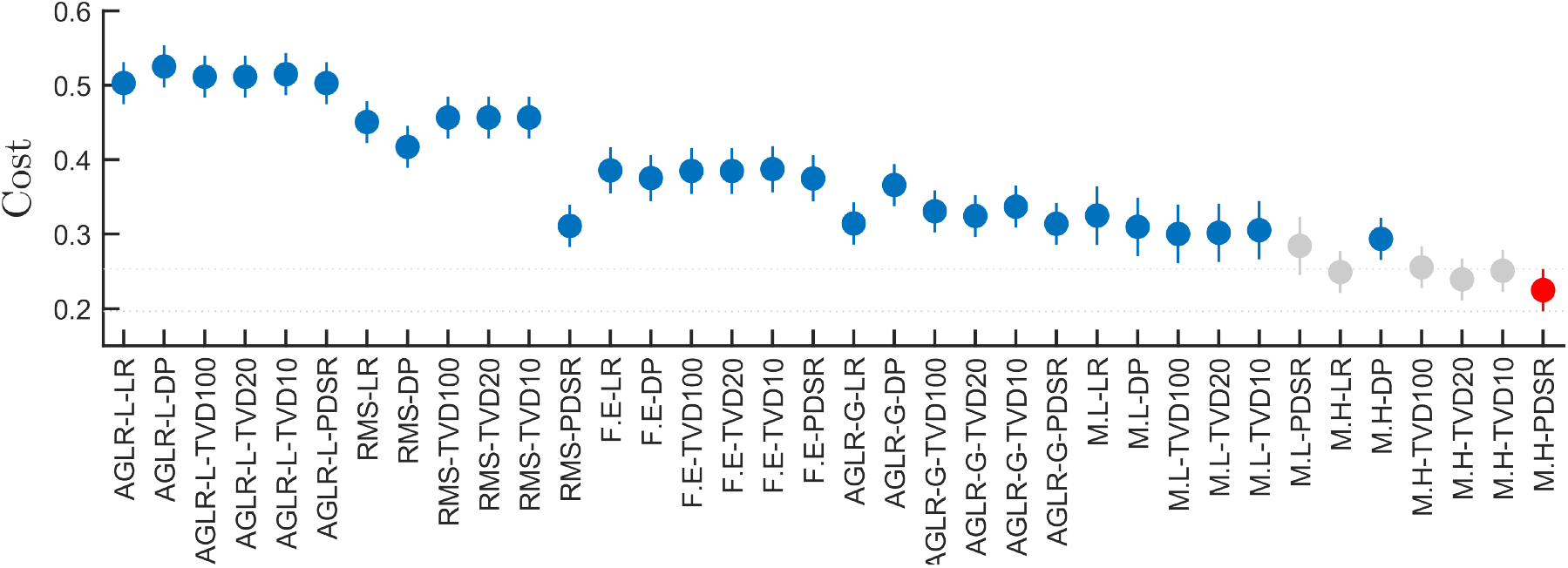
The error plot of cost of detection of the different detectors-separation measure pairs is presented. The maximally separating modified Hodges detector trained using the separation measure PDSR is the combination that incurs the least cost (Highlighted in red). Grey plots are statistically insignificant from MH-PDSR pair

**Figure 3.3:**
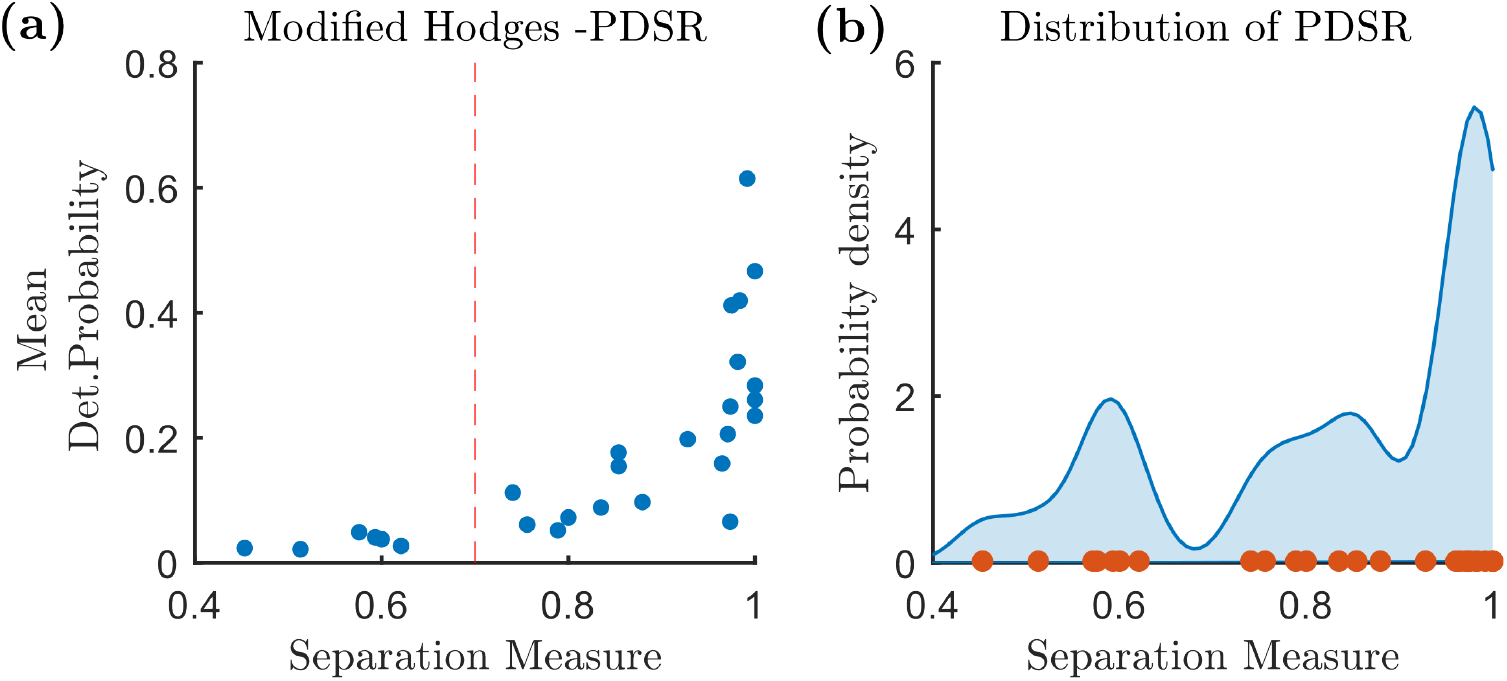
(a) Scatter plot of the optimum separation measure PDSR and the mean detection probability 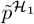 . The correlation coefficient is 0.7208 indicating a positive correlation between the two factors. (b) The kernel density plot of the PDSR separation measure with huge dip around 0.7

## 4 Discussion

Two questions need to be addressed to implement EMG-driven robot-assisted therapy for a severely impaired patient: (a) **Q1**: does the patient have sufficient residual EMG to drive robotic assistance during training? and (b) **Q2**: what is the optimal detector (high accuracy and low latency) to enable naturalistic human-machine interaction, if the patient has sufficient residual EMG? The work presented in this paper offers a practical approach to answering these questions using only data recorded from a patient under two conditions, while the patient: (a) is relaxed (i.e., no muscle activity ℋ_0_), and (b) attempt movements of interest (potential muscle activity ℋ_1_). The proposed unsupervised approach is based on the idea of maximally separating data under ℋ_0_ and ℋ_1_, without requiring any additional expert/clinician-derived annotation or information. Among the six different detectors and separation measures investigated, the AGLR detector exhibited the highest separation for most separate measures, followed by the modified Hodges detector and modified Lidierth detector. However, when evaluated using clinician-marked ground truth, the modified Hodges detector optimized using the PDSR measure had the lowest detection cost; The modified Lidierth-PDSR pair also incurs a cost that is statistically insignificant compared to the modified Hodges-PDSR pair. The modified Hodges detector had consistently lower detection cost than the other detectors. Interestingly, we found that the PDSR measure can also be used to screen severely impaired patients for the presence/absence of sufficient residual EMG to undergo EMG-driven robot-assisted therapy. Based on these results, a practical approach to answering questions Q1 and Q2 is to find the modified Hodges detector that maximally separates a patient’s data under ℋ_0_ and ℋ_1_ using the PDSR separation measure. If the PDSR value is greater than 0.7, then the patient has residual EMG (Q1) and the identified maximally separating modified Hodges detector is the optimal detector EMG detector of interest (Q2). If the PDSR value is less than 0.7 then the patient is unsuitable for EMG-drive robot-assisted therapy.

To our knowledge, this is the first study investigating an unsupervised approach to identify a patient-specific EMG detector when the presence/absence residual EMG and its exact times of occurrence are unknown. Most prior work have employed a supervised learning approach where the EMG epochs and their onset times are known (15; 16; 17; 12; 18); either because the data was simulated (13) or expert-marked annotation was available for the real EMG data (18). Such supervised approaches were useful to understand the detection properties of EMG detectors, but are impractical for routine use; as it is not possible to have expert annotation for each potential patient to be screened. Interestingly, we observed some interesting similarities and differences between the outcomes of the unsupervised approaches in this study and the supervised approaches employed previously (14). Staude et. al. observed that the AGLR detector to be well suited for EMG onset detection using simulated EMG data (13). The AGLR detectors were also found to be suitable for detecting EMG accurately with low latency in our previous work using simulated EMG (14). These previous observations appear to be in agreement with the current study where the AGLR detectors were found to have the best separation between the data under ℋ_0_ and ℋ_1_ across the study population and for most separation measures. Surprisingly, when evaluated on a data with ground-truth, the AGLR detectors do not offer the best detection performance. The AGLR detectors were optimized for detecting EMG onset events using a model for the EMG data that has a step increase in its amplitude at an onset time. The real data employed in this study does not conform to this model, which could explains its suboptimal performance. The real EMG signal from patients occurs in bouts of unknown onset times and duration, the signal amplitude during these bouts are not fixed, and the signal bouts have smooth onsets and offsets. Apart from the AGLR detectors, our previous work also identified the Fuzzy entropy detector and the modified Hodges detector to have consistently good performance (14). The Fuzzy entropy detector had the worst separation performance among the six detectors. The reason for this is unclear. The modified Hodges detector had consistently good performance with both supervised and unsupervised approaches which may be due to its simple structure that requires minimal assumptions about the signal model. Interestingly the modified Hodges detector appears to be least sensitive to separation measure employed for optimization.

The PDSR measure was found to have the lowest detection cost among the six separation measures. The TVD measures were sensitive to the number of bins used for the histogram, which was the reason, three different bin numbers were tried in the current study. The PDSR measure is a compromise between the DP and LR measures, which rewards large positive differences between the 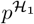 and 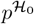, while penalizing large values for 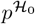 . The DP does not penalize for large absolute values for 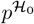. The LR penalizes large values for 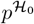, but does not reward positive differences between 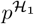 and 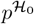 . The balanced nature of the PDSR measure could be the reason for it superior performance compared to the other measures.

The threshold of 0.7 was estimated for the PDSR measure for the maximally separating modified Hodges detector using the dataset employed in the current study. The kernel density of the PDSR separation measure shows a dip around 0.7, and the probability of detection 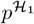appears to rise sharply above 0.7. This threshold of 0.7 corresponds to the following relationship between 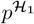 and 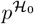,

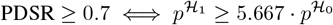

Assuming that the maximally separating modified Hodges detector is a good EMG presence estimator, patients producing EMG while attempting movements at a detection rate 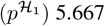 times the false positive rate 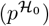 are deemed to have residual EMG that can be consistently detected. We identified 24 patients with separation values greater as seen in Fig.3.3. In our previous work employing an RMS detector, 22 patients were categorized as having sufficient residual EMG. Note that the RMS detector does not perform as well as the modified Hodges detector, and it was not optimized in our previous study (8). The use of an optimized detector that maximally separates data under ℋ_0_ and ℋ_1_ could be the reason behind the slight differences between the studies.

To evaluate the generalizability of the proposed unsupervised approach, its practical implementation, and it’s clinical utility, the following limitations need to be addressed :

1. We used an AR model in the current study to generate the data under ℋ_0_ for each subject and each trial. Ideally, this should be done with actual data recorded from the human subject while they are fully relaxed. However, appropriate AR models can reasonably approximate the spectral characteristic of EMG (19; 20). Real-world screening procedures must collect equal amount of data under both the relaxed (ℋ_0_) and movement-attempt (ℋ_1_) conditions to answer questions Q1 and Q2.
2. The study results are based on a small sample size of 30 chronic, severely impaired stroke subjects. The promising results of the current study warrant the investigation of the proposed unsupervised approach on a larger sample of severely impaired stroke subjects with no residual movements. Our current work is focused on carrying out a large screening study in the severely impaired stroke population, where equal amount of data is collected both under ℋ_0_ and ℋ_1_.
3. We were only focused on detecting EMG from a single channel of recoded data (EMG recorded from the two extensor muscles were considered as individual trials in the dataset). However, with the increasing availability of EMG arrays, multichannel EMG data can be easily employed for the EMG detection problem. Logical combinations (ANDing or ORing) of individually optimized detectors for each EMG channel might not be the best use of this rich multichannel data. Future work must investigate the best method to extend the idea of maximally separating detectors using multichannel data.
4. The current work does not shed any light on the expected quality of the human-machine interaction using the maximally separating detector. Experiments with human subjects on their sense of agency is required to evaluate the effectiveness of these subject-specific maximally separating detectors in implementing a natural human-machine interaction.

## 5 Conclusion

This paper presented an unsupervised approach to identify an optimal detector for EMG-driven robot assisted therapy using muscle activity recorded from patients while attempting movements. The proposed approach identifies a detector that maximally separates the data under relaxed and movement-attempt conditions - the *maximally separating detector*. The measure of separation in the data under these two conditions can be used as a screening tool for determining if a given patient has sufficient residual EMG to drive robot assisted therapy. The results identified the modified Hodges detector using the PDSR measure as the detector-separation measure combination that provides the best performance in terms of detection accuracy and latency. A separating value of 0.7 for the maximally separating modified Hodges detector using the PDSR measure was chosen as the threshold for screening if a subject has sufficient residual EMG. We believe that the proposed unsupervised approach holds value for quick screening of patients for the presence of residual EMG, and to identify the optimal detector to pick-up this signal. Future studies are needed to validate the findings of this study on a large sample of severe patients who cannot voluntarily move the hand, and to evaluate the quality of the human-machine interaction implemented using a maximally separating detector. We also note that this unsupervised approach can be easily applied to optimize detectors for other signal modalities including EEG.

## Data Availability

All data produced in the present study are available upon reasonable request to the authors

